# Early Detection Of COVID-19 Using A Smartwatch

**DOI:** 10.1101/2020.07.06.20147512

**Authors:** Tejaswini Mishra, Meng Wang, Ahmed A. Metwally, Gireesh K Bogu, Andrew W Brooks, Amir Bahmani, Arash Alavi, Alessandra Celli, Emily Higgs, Orit Dagan-Rosenfeld, Bethany Fay, Susan Kirkpatrick, Ryan Kellogg, Michelle Gibson, Tao Wang, Benjamin Rolnik, Ariel B Ganz, Xiao Li, Michael P Snyder

## Abstract

Wearable devices digitally measuring vital signs have been used for monitoring health and illness onset and have high potential for real-time monitoring and disease detection. As such they are potentially useful during public health crises, such as the current COVID-19 global pandemic. Using smartwatch data from 31 infected individuals identified from a cohort of over 5000 participants, we investigated the use of wearables for early, presymptomatic detection of COVID-19. From physiological and activity data, we first demonstrate that COVID-19 infections are associated with alterations in heart rate, steps and sleep in 80% of COVID-19 infection cases. Failure to detect these changes in the remaining patients often occurred in those with chronic respiratory/lung disease. Importantly the physiological alterations were detected prior to, or at, symptom onset in over 85% of the positive cases (21/24), in some cases nine or more days before symptoms. Through daily surveys we can track physiological changes with symptom onset and severity. Finally, we develop a method to detect onset of COVID-19 infection in real-time which detects 67% of infection cases at or before symptom onset. Our study provides a roadmap to a rapid and universal diagnostic method for the large-scale detection of respiratory viral infections in advance of symptoms, highlighting a useful approach for managing epidemics using digital tracking and health monitoring.

## Introduction

Early detection of infectious disease is important to mitigate disease spread through self-isolation and early and effective treatments. Presently most diagnostic methods involve sampling nasal fluids, saliva or blood followed by nucleic acid-based tests for detecting active infections or blood-based serological detection for past infections. Although highly sensitive, nucleic acid-based diagnostics may require samples gathered several days post-exposure for unambiguous positive detection ^1^. Moreover, they cannot be implemented routinely at low cost and are constrained by emerging shortages in key reagents.

Consumable wearable devices are an accurate and widely deployed platform from which to establish individual baseline parameters of health, which may be used to detect significant deviations from baseline physiology at the onset of infection. ^2–4^. We have previously shown that smartwatches and simple pulse oximeters can be used for active detection of Lyme disease, and, in retrospective studies, heart rate and skin temperature can be used to detect viral respiratory infections, including asymptomatic infections ^5^. Wearable sensors have also been used to detect atrial fibrillation ^6^. Other recent studies have shown that elevated heart rate measurements from smartwatches can be used in epidemiological studies to track the spread of respiratory viruses ^7, 8^.

The use of wearable devices has significant potential to mitigate the COVID-19 or Coronavirus Disease 2019 global pandemic. This pandemic has already infected 10.1 million individuals, causing 502,278 deaths worldwide ^9^. There is substantial need and opportunity for population-scale technology solutions for infection detection and tracking ^10^. Active infections are currently tracked using PCR assays, which may require up to three days post-infection for a reliable positive signal ^1^. In addition, the test is not generally utilized on a daily basis. Moreover, since most infections become apparent only upon symptom onset, the current methods of testing cannot identify presymptomatic carriers, which is a significant challenge for the implementation of early stage interventions that reduce transmission. It is believed that as much as 50% of COVID-19 cases are asymptomatic, facilitating further viral spread ^11,12^. As such, accessible and inexpensive methods for early detection of COVID-19 in real-time are urgently needed.

Smartwatches and other wearable devices are already used by tens of millions of users worldwide and measure many physiological parameters such as heart rate, skin temperature and sleep ^13^. Here, we investigate the use of wearable devices for early detection of COVID-19 in a retrospective manner, as well as an approach for using wearable-detected physiological parameters for real-time health monitoring and surveillance. Using heart rate (HR) and steps data from a large cohort of 5,322 individuals, we show that HR signals from fitness trackers can be used to retrospectively detect COVID-19 infection well in advance of symptom start (“offline detection”). In addition, we developed an “online detection” algorithm to identify early stages of infection by real-time monitoring of HR. We also examine the association between symptom type and severity, heart rate signals, and the effect of infection on activity and sleep.

## Results

### Study Design and Overview

We investigated whether smartwatches could be used to detect COVID-19 at an early, presymptomatic stage. The overall study design (Fig. 1A) was to enroll a cohort of participants who had a self-reported COVID-19 or other infections as well as a wearable device capable of detecting heart rate, steps and other physiological measurements. We then examined whether physiological deviations from baseline were detected around the period of illness, the detection frequency and timing of onset of the event, and associations with symptoms. Finally, we established an online method for potential real-time early detection of illness onset using retrospective data.

**Figure 1.**
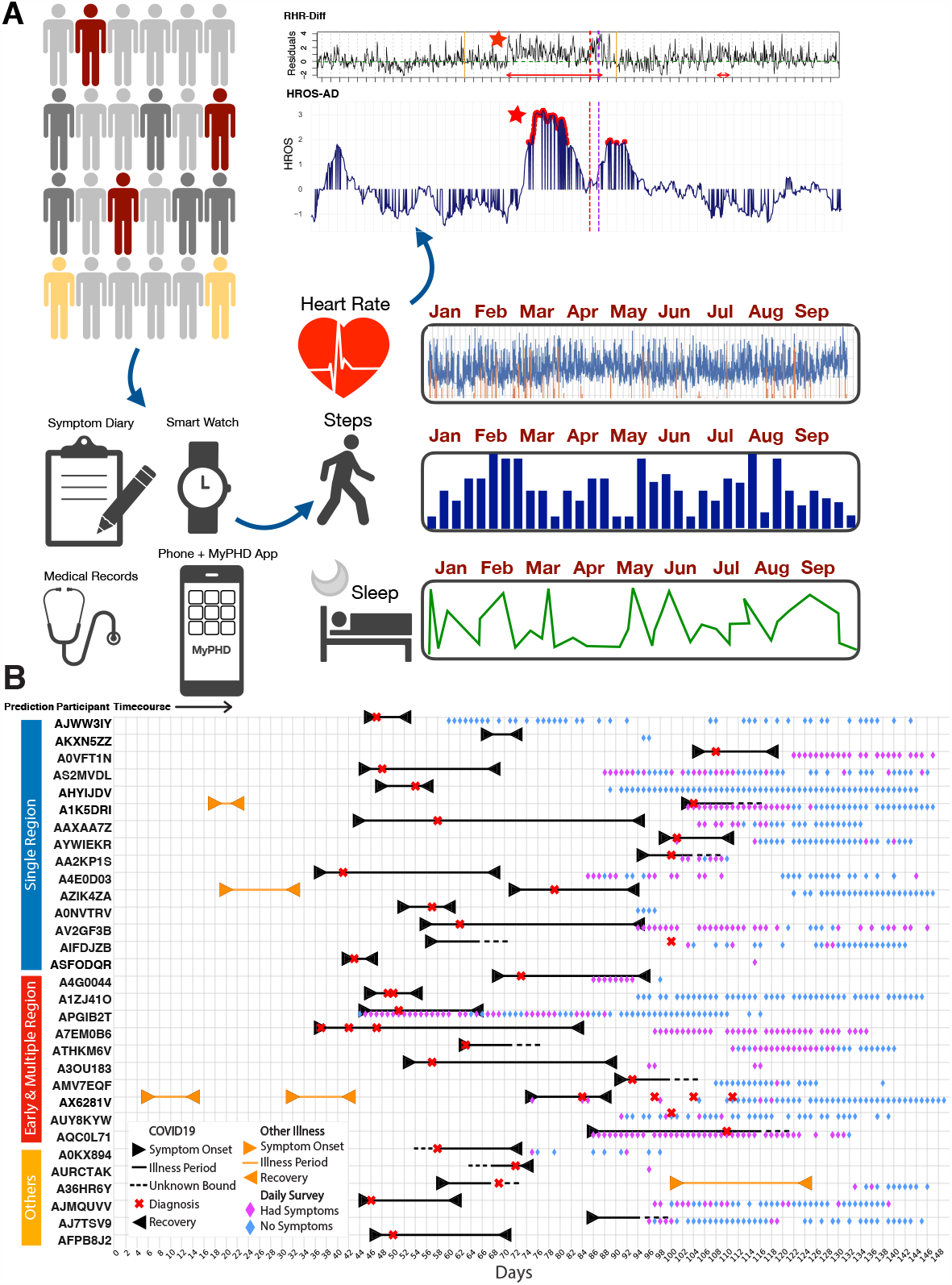
Overview of study design, cohort, and data. **(A)** Overview of study design. 5,262 participants were recruited, including individuals who were (i) sick and tested positive for COVID-19 (dark red), (ii) sick and tested positive for other illnesses (gold), (iii) sick without a confirmed diagnosis (dark grey), and (iv) participants who were not sick but are at high risk of exposure (light grey). Participants were asked to log daily symptoms, and to share their fitness tracker data via the study app, MyPHD. Data types collected include heart rate, steps and sleep over a period of several months. Two infection detection algorithms were developed (RHR-Diff and HROS-AD). Panels represent derived heart rate metrics (X-axis) over a period of months in one individual. (Y-axis = standardized heart rate residuals (top), and standardized heart rate (bottom), X-axis = days). The earliest detected abnormal heart rate elevations are marked by a red star.The detected anomaly periods are shown in red arrows in RHR-Diff. The anomaly time points are shown in red dots in HROS-AD. Symptom onset day and diagnosis day are indicated, (vertical dashed relines respectively), as is the infection detection window of −15 to +7 days before symptom onset (solid gold vertical lines) **(B)** Summary of data collected from 31 study participants who reported a confirmed diagnosis of COVID-19 with a symptom onset and/or test date. Each row along the Y-axis represents one participant labeled by prediction groups (details in Fig. 2), and columns along the X-axis represent days. The plot shows sick periods for COVID-19 (between black arrows, with dashed lines for unknown bounds where symptom onset or recovery day was unclear), COVID-19 test date (red x symbol), sick periods for other illness (between orange arrows), and the days that participants filled in the daily survey as purple diamonds representing days symptoms were reported and blue diamonds days when symptoms were not reported.

Under an IRB-approved study (Stanford IRB #55577) we enrolled 5,262 participants who completed surveys of illness, diagnosis and symptom dates, illness severity and symptom type (Fig. 1A, 1B, Table 1, Supplementary Tables 1 - 4) and collected data through a secure REDCap system. 4,642 of these participants reported wearing a smartwatch: 3,325 wore Fitbits, 984 wore Apple watches and 428 wore Garmin, and the remaining wore other devices (Fig. S1). Of these, 114 individuals reported a COVID-19 illness with symptom and diagnosis dates and another 47 individuals reported a different respiratory infection with symptom and diagnosis dates for an identified pathogen. Since most people wore Fitbits, we focused on this group (31 in total, Supplementary Table 1): 26 had smartwatch data spanning and adjacent to the COVID-19 disease dates as well as symptom dates and diagnosis dates. An additional 5 individuals with Fitbit devices lacked either a reported symptom date or diagnosis date. Of note many (n = 5) participants had wearable data but lacked measurements at or shortly after the time of infection, suggesting that many participants do not habitually wear their devices when ill (for e.g. participant *AV2GF3B* in Fig. S2).

**Table 1:**
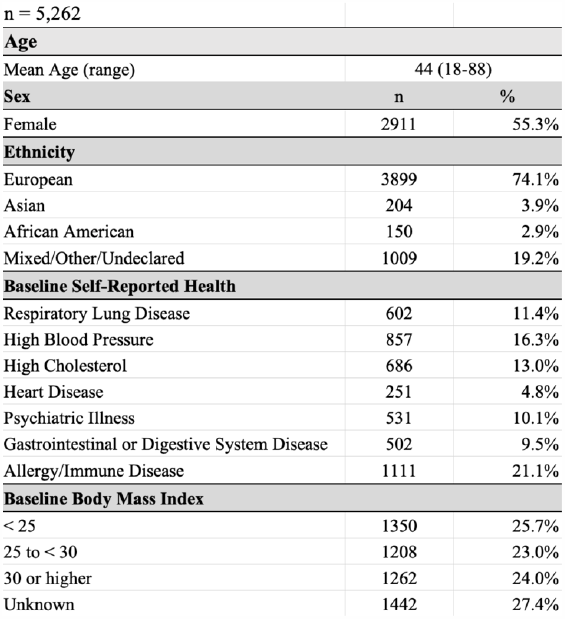
Demographics and Health Characteristics of the Cohort.

### Abnormal resting heart rate, heart rate/steps ratios and sleep are associated with COVID-19 illness

We first determined if abnormal physiological events are associated with SARS-CoV2 infection and if these can be detected using a smartwatch at or near the time of COVID-19 infection. We focused on the Fitbit data, which had the highest number of complete datasets. Three parameters were investigated: elevated resting heart rate (RHR) relative to a previous “healthy window”, an increased heart rate relative to step (i.e. resting heart rate/steps ratio), and sleep (see Methods). We focused primarily on early onset events since participants often take medications or undergo other treatments once symptomatic.

We developed two methods for detecting aberrant physiology: 1) In the RHR Difference (RHR-Diff) method, we detect and identify elevated RHR time intervals based on the standardized residuals (see Methods). The standardized residuals were constructed at one-hour resolution by comparing each interval to the average daily curve using a 28-days sliding window. We applied a nonparametric approach ^14^ to test whether the sequence of standardized residuals is a homogeneous process. Under the significance level 0.05, the elevated regions were reported as abnormal RHR periods (Supplementary Table 5). 2). In the anomaly detection method (HROS-AD), we created a new feature known as HROS (Heart Rate Over Steps) by dividing heart rate with steps data and compare HROS at each hourly interval with the rest of the intervals using Gaussian density estimation ^15,16^. Prior to the comparison, we smoothed and standardized the HROS (see Methods). Using Gaussian density estimation, we computed an anomaly score for each observation and classified them as normal (1) or anomaly (−1) with an outlier threshold of 0.1 (see Methods, Supplementary Table 6). Analyses were performed on all the data available for each individual, including data collected before, during, and after the reported illness. A method similar to HROS-AD, RHR-AD (based on resting heart rate, see Methods) produced similar results (Supplementary Tables 7, 10).

Using dates of symptom onset and diagnosis to define sick periods, we then defined a sickness detection window for each individual based on symptom onset date wherever available (14 days prior to 7 days after), and diagnosis date when symptom date was not available (three cases). We scored both of our detection methods based on the interval (RHR-Diff) or HROS-AD period that overlapped with the sickness detection window. Of 31 individuals, we found outlying periods near the time of infection using either RHR-Diff or HROS-AD in 25 (81%) with 23 identified by both methods (Supplementary Tables 8 - 9, Supplementary Table 10 for RHR-AD results). RHR-Diff detects one high signal region not identified by HROS-AD and, similarly, HROS-AD detects one high signal region not identified by RHR-Diff. Strikingly, we observed that neither method detected stable signal regions specifically at COVID-19 infected regions in six individuals as described below.

The 31 individuals fell into three types of patterns (Fig. 2A). Group I is a set of 15 individuals for whom we were able to detect disease primarily as a single elevated period or a tight cluster of elevated periods near or overlapping the disease period. Two examples are shown in Fig. 2B and 2C in which the heart rates are elevated starting 15 and 4 days prior to symptom onset and 16 and 10 days prior to diagnosis, respectively. In cases where a tight cluster of elevated periods is observed, it is possible that the normal physiological periods reflect times of remission or medication.

**Figure 2.**
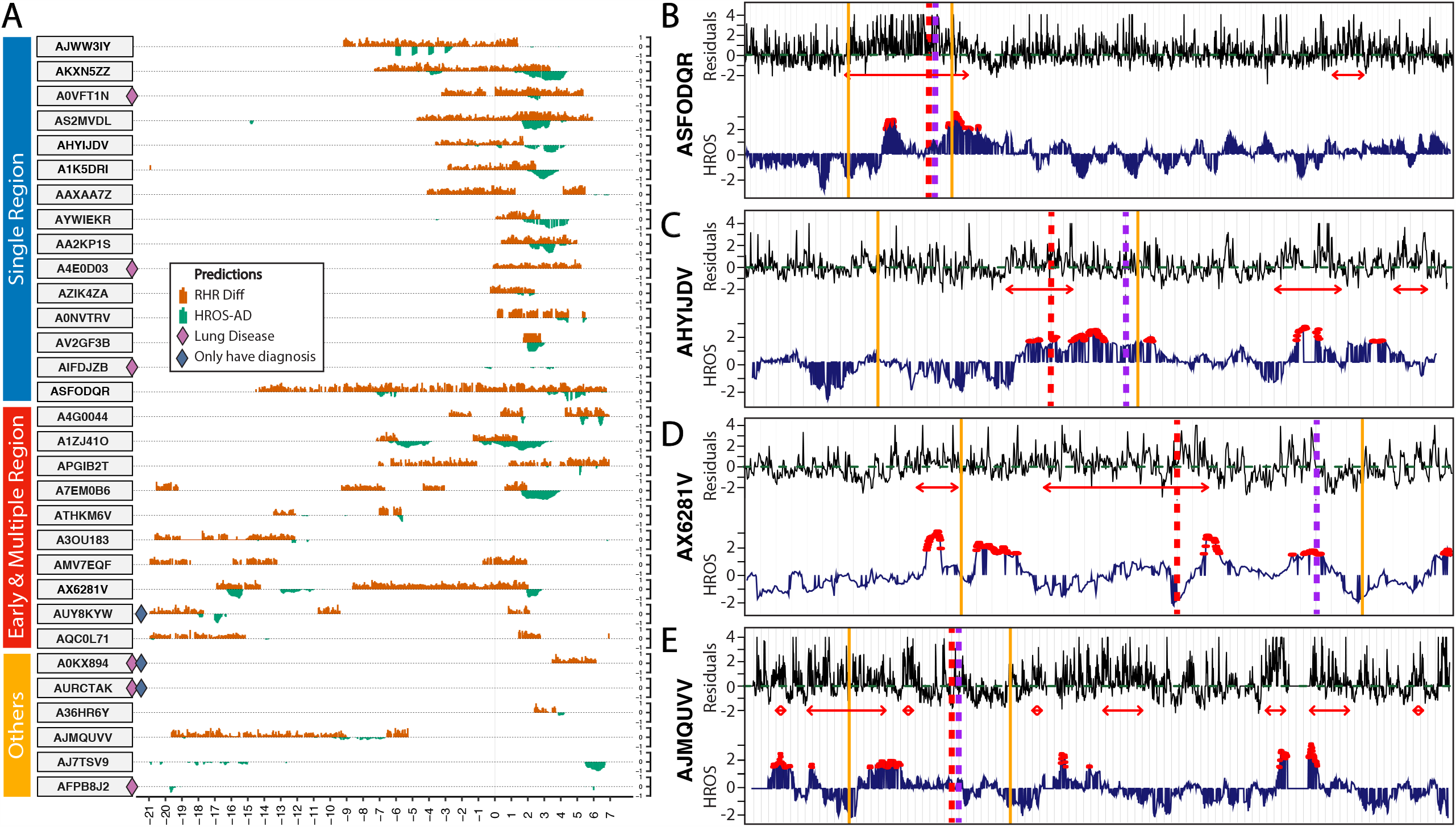
Association of heart rate with COVID-19 illness. **(A)** Two-sided bar plots depicting heart rate metrics and timing of infection detection from RHR-Diff and HROS-AD with respect to the infection detection window for the 31 participants. The plots are manually grouped into three groups, “Single Region” (blue), “Early & Multiple Region” (red), and “Other” (gold). X-axis is days during the infection detection window, and Y-axis is standardized and rescaled (0 to 1) residual values from RHR-Diff (coral bars), and standardized and rescaled (0 to −1) HROS values from HROS-AD (inverted green bars) in the intervals during which COVID-19 infection was detected by each algorithm. These values are plotted separately for each participant. This window is a period of time centered around symptom onset, Day 0 (substituted by diagnosis day wherever day of symptom onset was unavailable). The infection detection window spans a period of −14 days before Day 0 and +7 days after Day 0. Note that participant *A3OU183* had a separate detection event after the COVID-19 illness that is not shown in this plot. **(B) - (E)** Examples of heart rate metrics during COVID-19 infection for four participants, two from Group I (B and C), and one each from Group II (D), and Group III (E). For each pair of plots, the X-axis is the number of days (each tick is a day), and the Y-axis is the heart rate metric. Red and purple vertical dashed lines indicate day of symptom onset and diagnosis, respectively, for both panels. Shown are standardized Heart Rate Over Steps or HROS from the HROS-AD method (bottom panel, dark blue line), and standardized heart rate residuals from the RHR-Diff method (top panel, black line). For RHR-Diff, the green dashed line is at zero. Yellow/gold vertical solid lines mark the infection detection window used to score detections as a hit or a miss. Also indicated are time intervals where the heart rate residuals are significantly elevated from RHR-Diff (red arrows in top panel), and times when anomalies are detected by HROS-AD (red dots, bottom panel).

Group II had 10 individuals and formed a cluster where we were able to detect a symptom associated peak as well as an earlier significant elevation period within 28 days of the symptom onset based on RHR-Diff (+7/−21 days). An example is shown in Fig. 2D. In some cases this affected the ability to clearly discern the onset of COVID-19 physiology period. Three of these individuals had a self-reported stress period (either illness or other), raising the possibility that the stress-associated event may have contributed to COVID-19 illness onset.

Group III consisted of the six individuals for whom a single stable elevated period could not be easily discerned at symptom onset or early diagnosis; these individuals often had many signals distributed across a substantial period of time (Fig. 2E is an example) or no significant signal.

Interestingly, three of these individuals had respiratory lung disease and often required medication. It is likely that these conditions interfered with outlier detection. However, not all individuals with respiratory conditions are missed using a Fitbit; three other individuals with these conditions gave high RHR-Diff (Fig. S2) and HROS (Fig. S3) signals associated with illness.

For Groups I and II the number of days between the beginning of the aberrant signal and date of symptom onset (when available), and date of diagnosis are distributed as shown in Fig. 3A and 3B (Supplementary Table 11). 87.5% (21 out of 24) and 100% (24 out of 24) of cases show elevated signals in advance or at the time of symptom onset or diagnosis, respectively, and the median values are 3 days and 7 days in advance, respectively. The median heart rate increase in the first period of onset is 7 beats/min with a broad range (Fig. 3C, Supplementary Table 12). Overall, these results indicate that altered physiology is associated with COVID-19 illness, often in advance of symptoms and that this can be detected with a wearable device.

**Figure 3.**
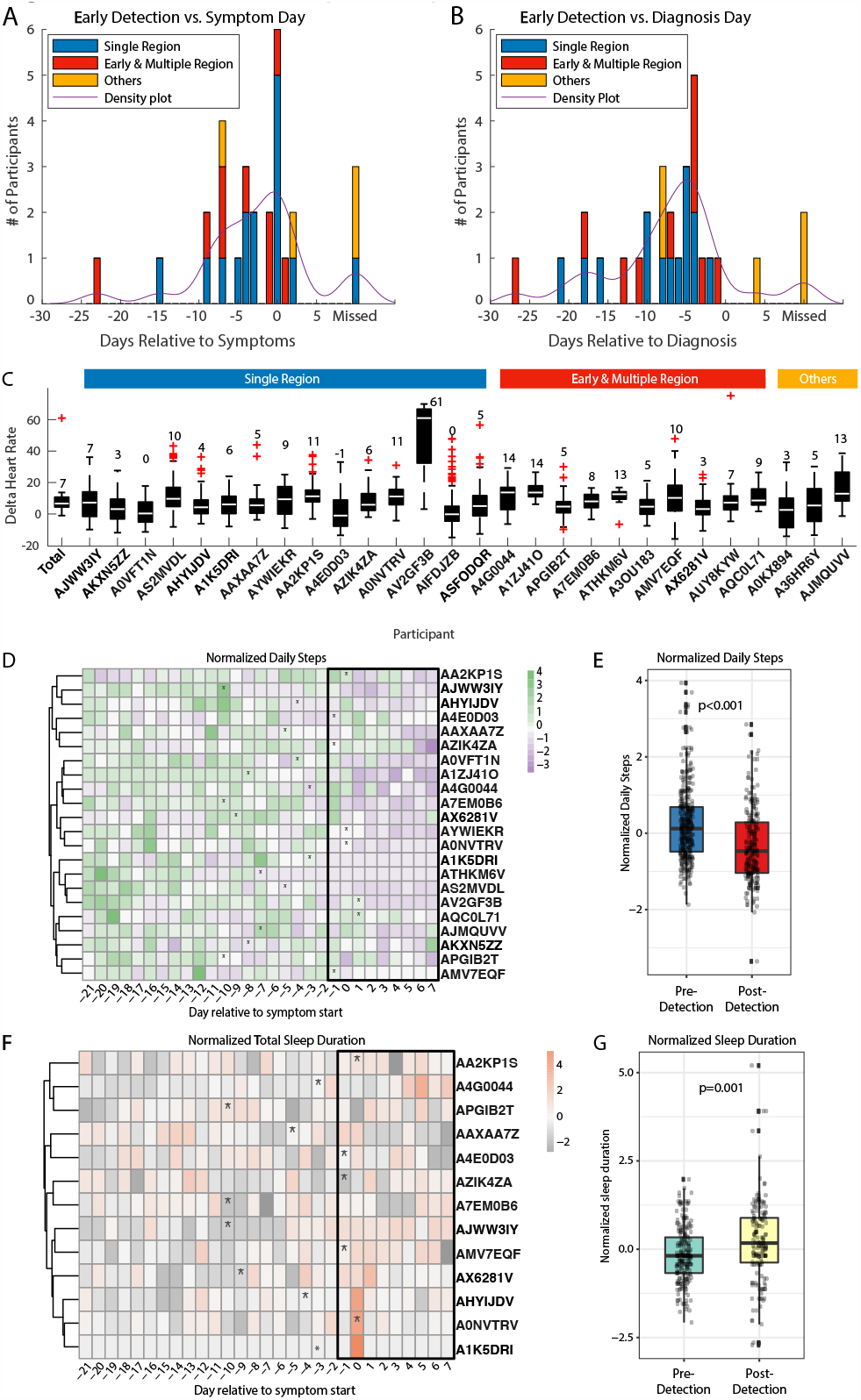
Summary of detection timing, heart rate, steps and sleep during COVID-19 illness. **(A), (B)** Histogram summarizing the distribution of the early detection of COVID-19 events compared to the first day of self-reported symptoms **(A)** or the reported diagnosis day **(B)**. If multiple RHR-diff intervals exist within or intersecting the COVID-19 event (+7/−14 days of the symptom day), the first day of the closet interval will be used. If the symptom day is not available or there is no interval detected within the 21 days of the symptom onsets, we will take the closest interval within the 28 days of the diagnosis onset (+7/−14 days). The colors represent the number of individuals in each group, Group I (blue), II (red), and III (gold). The purple line shows the kernel probability density estimate. Also shown are individuals for whom the algorithm missed detecting COVID-19 infection. **(C)** Box plot summarizing the delta resting heart rate (RHR) of the detected COVID-19 interval compared to the baseline RHR of the same individual. This box plot excludes individuals for whom the RHR-Diff missed. The number above each box plot represents the median value of the RHR. **(D)** Heatmap showing standardized daily steps per participant (i.e. z-scores of daily steps), for the 22 participants we have steps data for and for whom our RHR-diff detects change of RHR any day between 14 days prior to the symptoms date and 2 days after. X-axis represents the day relative to the reported start of symptoms date, Y-axis represents participants, asterisk represents the first day of detection. **(E)** Boxplot showing change in daily steps between days before and after the detection start date in a window of −21 days before to +7 days after symptoms onset date. **(F)** Heatmap showing standardized sleep duration per participant, for the 13 participants we have sleep data for and for whom our RHR-diff detects change of RHR any day between 14 days prior to the symptoms date and 2 days after. **(G)** Boxplot showing change in total sleep duration between days before and after the detection start date in a window of −21 days before to +7 days after symptoms onset date.

### Sleep and activity alterations associated with COVID-19 illness

Having established aberrant physiological signals associated with COVID-19 illness, we investigated whether COVID-19 also affected behavior, specifically steps and sleep duration (Fig. 3D - G, Supplementary Tables 13 – 18, see Methods). Although Fitbit devices are not considered gold standards for many sleep parameters, they are widely used. We examined the parameters reported by the manufacturer (see Methods) and found that steps significantly decreased at the onset of the outlying RHR-Diff signal associated with COVID-19 illness (linear mixed model (LMM), p < 0.001; Fig. 3E, S4A). Sleep duration significantly increased after the onset of the outlying RHR-Diff signal (LMM, p = 0.001, Fig. 3G, S4B). These results indicate that COVID-19 illness alters steps and sleep patterns, which can be tracked using a wearable device. Interestingly, many of these are detected prior to symptom onset.

### Association between heart rate signals and symptoms

A subset of participants filled out daily logs before or during their COVID-19 illness, providing a detailed time course of symptom severity, progression, and relapse (Fig. 4A-D), while others filled out detailed past illness surveys that summarized their symptoms over the entire illness period (Fig. 4E). The first individual (Fig. 4A) began daily logs when symptoms developed, reporting a 22-day period of mild-to-moderate coughing, fatigue, and aches and pains that was anticipated by both the RHR-Diff and HROS-AD heart rate metrics. Symptoms then deteriorated rapidly coupled with abnormal physiological signals suggested by both algorithms elevated temperature, and positive COVID-19 test. The participant was admitted to the hospital 5 days later and it was a total of 41 days from symptom onset to recovery. A second participant (Fig. 4B) had an early RHR-Diff signal a week before symptom onset. The disease progressed quickly into severe diarrhea, fatigue, headaches, elevated temperature and positive COVID-19 peaking in severity and then declining over two weeks. In total 18 days of initial illness were followed by 12 days where the participant felt recovered, before a relapse characterized by elevated temperature, fatigue, diarrhea, and elevated heart metric signals. A third participant reported COVID-19 illness lasting 13 days (Fig. 4C) that was led by a RHR Difference alarm, followed by ongoing symptoms of fatigue and occasional chest pains. Heart rate metrics alarms accompanied the return of shortness of breath, for which the participant was hospitalized 35 days after initial symptom onset. Daily logs began at symptom onset for the fourth participant (Fig. 4D), three days after an RHR-Diff alarm. Illness progressed over 23 days with a rapid rise in temperature and HROS-AD alarms accompanied by severe fatigue, aches and pains, and slow recovery.

**Figure 4.**
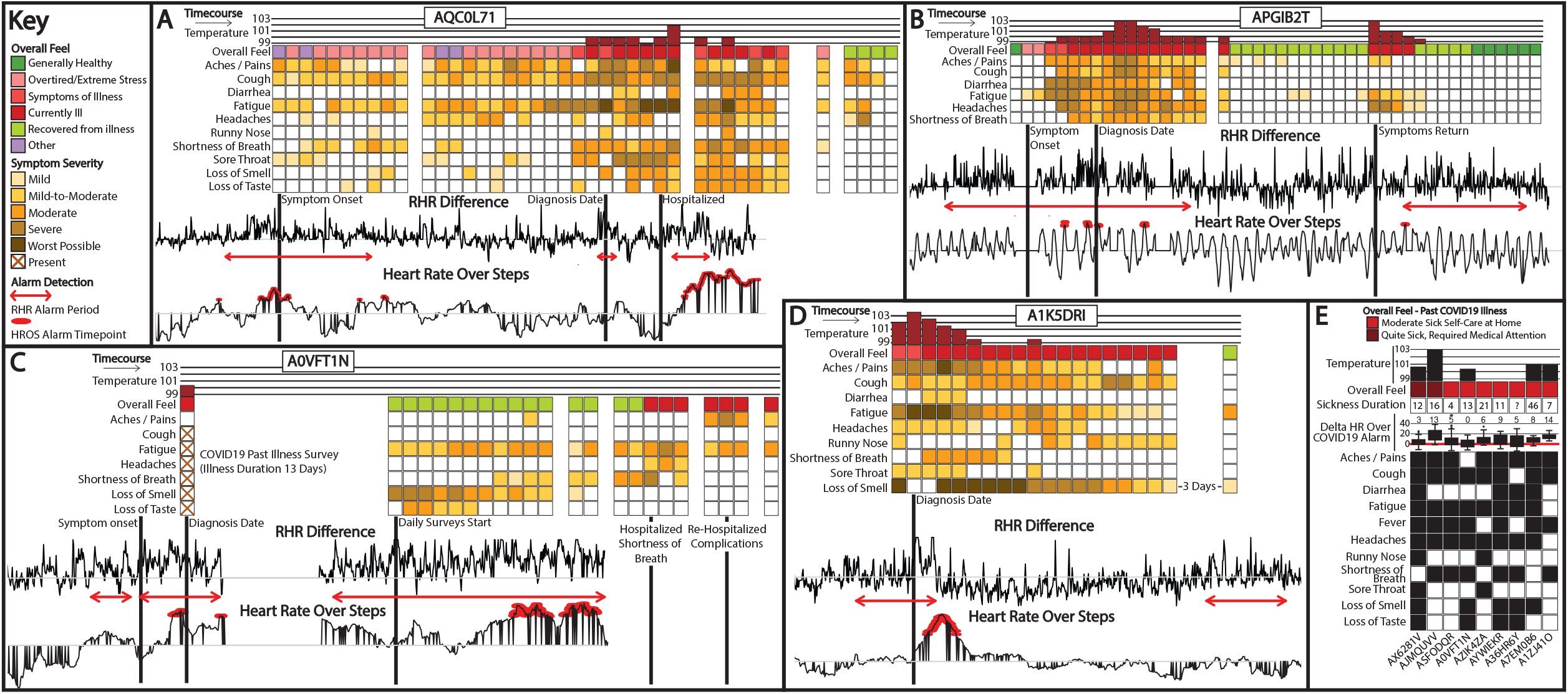
Association of COVID-19 symptoms with heart rate signal. **(A-D)** Plots of four individual participants over the course of COVID-19 infection. Vertical columns along the y-axis each represent a single day of symptoms aligned with heart rate metrics below, and days progress along the x-axis from early illness to late. Columns of symptom boxes are only present on days the daily survey was completed, while heart rate metrics progress continually below. Overall feel indicates how the participants reported feeling on a particular day, with a barplot above indicating measured temperature if reported, and specific symptoms highlighted below as a heatmap depicting severity below. Black vertical lines below symptoms and descending into heart rate metrics are labeled to highlight significant days during the illness course and align with symptoms above. The RHR Difference panel shows standardized heart rate residuals from RHR-Diff (black lines), and time intervals where the heart rate residuals are significantly elevated (red line and arrows). The bottom panel shows standardized Heart Rate Over Steps using the HROS-AD method (black line), and each detected anomaly is indicated by a red oval. **(E)** Symptoms for individuals who provided surveys on a past COVID-19 illness. Each column represents a study participant labeled at the bottom. Starting at the top of each column is a barplot if temperature was reported during the illness, followed below by overall feel according to the two colors above, the total duration between reported symptom onset and recovery if provided, a boxplot and the numerical median of delta RHR were when heart rate residual alarms are raised, and then boxes for individual symptoms with black boxes indicating reported symptoms and white for symptoms not reported.

Lastly, in addition to the daily survey we also examined symptoms reported post illness i.e. retrospectively. In this limited sample, we did not detect any obvious association in magnitude of RHR differences during alarm periods with symptom type, number, illness length, or temperature (Fig. 4E). Overall, at the individual level, COVID-19 progression and severity are generally concordant with heart rate metrics, but these cases highlight temporal and individual variation more widely observed with the illness ^12,17^.

### An approach to detect early COVID-19 onset in real-time

The ability to detect altered physiology in advance of symptoms raises the possibility that a method can be developed to detect early stages of COVID-19 illness in advance of symptoms using a smartwatch. To test this possibility, we developed an “online” detection method called CuSum (see Methods). This detection was based on a type of cumulative statistics ^18,19,20^ which cumulates the deviations of the elevated residual RHRs. The test statistics from the baseline records in the previous 28 day sliding window build an empirical null distribution. We report a warning alarm as the first time we observe a test statistic more extreme compared to the null, with a p-value generated from comparing current test statistics to the measurements in the baseline. To reduce the number of alarms, a two-tiered warning system was developed. An initial warning alarm (yellow light) signals when the first time the p-value was less than 0.01 (usually in the first few hours). Monitoring continues, and if it remains elevated over 24 hours, it signals a positive event (red alert; see Methods).

We tested this method initially on four individuals for whom we had collected >6 months of wearable data (Fig. 5A, 5B, Supplementary Table 19). In addition to the annotated COVID-19 infection, other strong elevated signals are identified as well as smaller signals. Some of these correspond to annotated infections. Others are not annotated but occur at periods that might be associated with increased heart rate. For example, three of the four individuals had high heart rate in the November-December holiday period which is commonly associated with air travel, alcohol, stress, as well as illness. A number of alarms of lower duration or signal are also observed.

**Figure 5.**
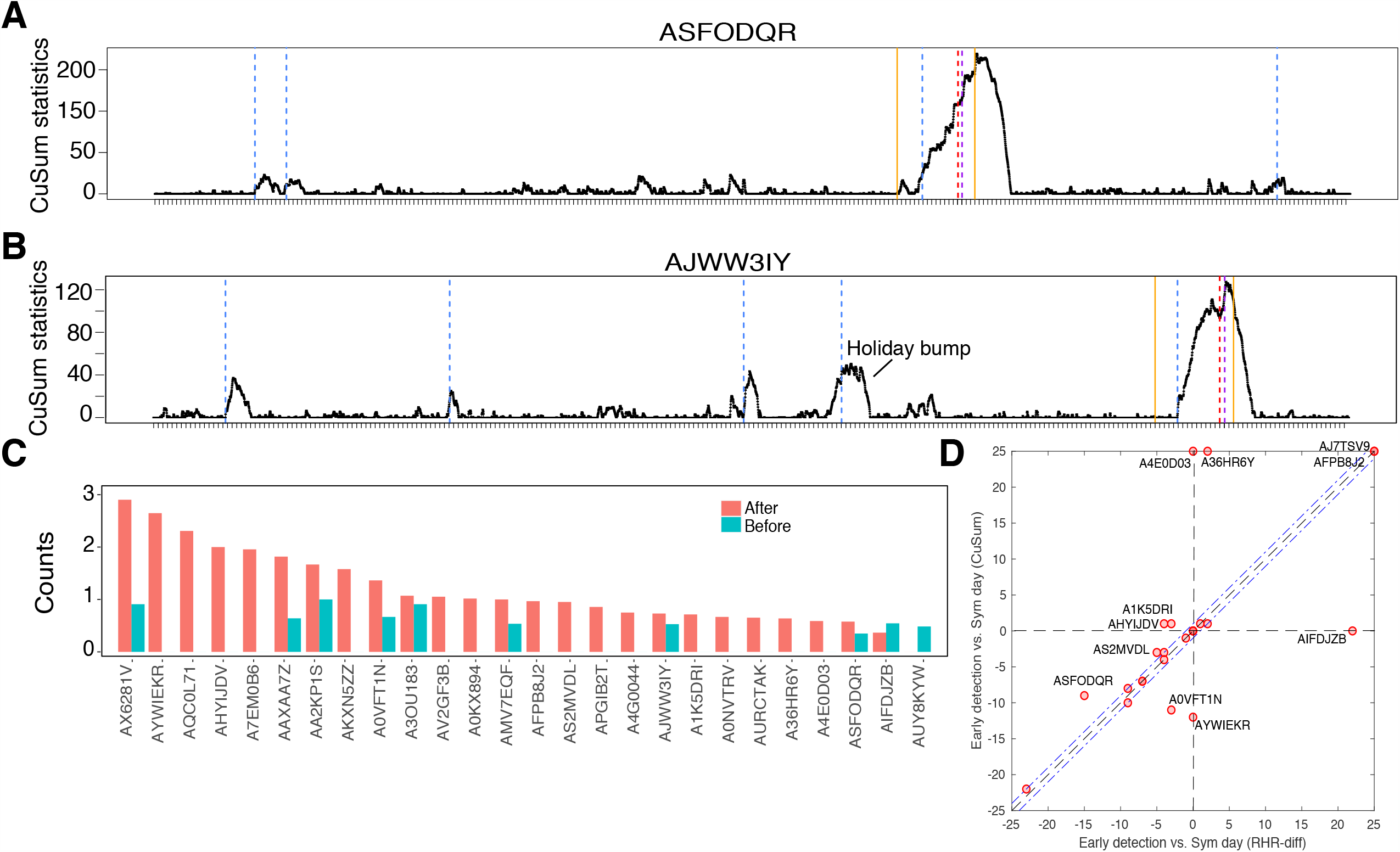
Online detection of COVID-19 infection. **(A-B)** Examples of online prediction performance during COVID-19 infection for two participants. For each pair of plots, the X-axis is the number of days (each tick is a day), and the Y-axis is the CuSum statistic. Red and purple vertical dashed lines indicate day of symptom onset and diagnosis, respectively. The blue vertical lines indicate the alarming time from the online detection (see Methods). **(C)** Alarm counts per 30 days for participants. The blue and red bars indicate the alarm counts before and after the COVID-19 event. Average alarm counts are 0.25 vs 1.19 before and after. **(D)** Early detection comparison between offline detection (RHR-Diff) and online detection (CuSum). Detection days are compared to the symptom day. Each red circle dot indicates one participant.

We also examined all 24 individuals who had at least 28 days of data ahead of symptom onset (Supplementary Table 20). 67% (16/24) had an alarm on or before COVID-19 symptom onset using the CuSum alarm model. Four more people were late for one day compared to the first day of the self-reported symptom. Of the remainder, two were people who were previously missed in offline detection and three had respiratory illnesses that were difficult to detect in our initial retrospective study. As shown in Fig. 5C, ten individuals had non-COVID-19 alarms prior to COVID-19 infections ranging from 0.35 to 1 alarms per month. Interestingly the number of alarms increases considerably post-COVID-19 infection, suggesting lingering complications from COVID-19 illness.

We compared our online detection results with those observed using the RHR-Diff approach and in general found good agreement (less than one day difference or both methods were missed) in 15 of the cases (Fig. 5D, Supplementary Table 21). However, 3 cases were detected earlier using the online CuSum approach, and 4 cases were detected late, and 2 cases were missed only in the CuSum version. 3 out of 4 cases missed by the CuSum have preexisting conditions (e.g, respiratory lung disease, psychiatric disease). Other differences are often due to the difference between the online and offline difference (see Fig. S2 for examples). RHR-diff detect significant intervals based on the global dataset while the detection of CuSum are solely based on the data received upfront.

## Discussion

From a sizable cohort, we identified a number of individuals who tested positive for COVID-19 and other illnesses and wore a smartwatch. Using these data, we found that elevated resting heart rates and outlying HR/steps measurements were altered, usually in advance of the symptoms. The early times of detection are generally consistent with the latent period of presymptomatic illness reported previously ^11^. In two Group I individuals the signal was observed 9 days or earlier. Because the actual timing of infection in these cases is not known, it is possible that these and other events represent early stress events that merge into the COVID-19 illness (e.g. Fig. 4A). Indeed, in 10 Group II individuals a discrete early event is observed and in three individuals this has been associated with a self-reported illness or family stress event. It is likely that early stress events increase vulnerability to SARS-COV2, resulting in illness.

We used the information learned from the retrospective analysis to design a potential approach for real-time early detection of COVID-19 illness (CuSum). In addition to detection of the COVID-19 events, other events are identified, of which some likely reflect illnesses, as we observed previously ^5^. Many of the other events could reflect situations that stimulate sustained increased heart rate such as medication, alcohol, travel, emotional or other stress inducers. Indeed, three of four cases with data covering the holidays showed significant elevation of long duration. Those short in duration (e.g. due to watching a scary movie) will likely go off after a brief period of time. Thus, using our proposed two-tiered continuous alarm system, early events can be acted upon by self-isolation or, if an increased signal ensues, this can result in continued isolation, direct viral diagnostics and/or potentially consulting a physician.

It should be noted that these wearable devices are not yet FDA-approved and our study is still modest in size. Another limitation we observed is that some individuals do not wear their devices (or let their charge expire) when symptomatic, which may affect monitoring patterns. Patterns of non-use were observed both with Fitbit and for different devices and we expect that devices which require daily charging will also have more missing data. Nonetheless, devices whose charge lasts for several days should be powerful for early detection prior to loss of device function.

It is currently unclear as to whether our approach can distinguish infections from SARS-COV2 from those caused by other illnesses. A challenge is that COVID-19 has heterogeneous physiologic presentation between individuals ^21,22^, as observed in our study. Regardless, any illness onset information is valuable, especially during a pandemic and can be followed up with appropriate testing. It is also likely that other types of physiological measurements (e.g. heart rate variability, respiration rate, skin temperature, blood oxygen saturation, electrocardiogram) obtainable from wearable devices will be valuable for both distinguishing illnesses from different infectious agents and could be used to increase diagnostic sensitivity and perhaps even predict illness severity and symptoms ^23–26^. Data from reported respiratory rates and blood oxygen is expected to be particularly useful in COVID-19 prediction ^27^. At the time of this writing, such data was not available to us; however, such data, along with increased participant size, will greatly improve diagnostics. Regardless, this continuous monitoring approach is expected to be powerful for early infectious illness detection and offers many advantages over PCR as well as other biochemical based methods. Such information will inform patients for self-isolation, diagnosis confirmation, and early treatment.

## Methods

### Participant recruitment

We recruited 5,262 adult individuals for this study under protocol 55577 approved by the Stanford University Institutional Review Board. Participants were recruited using REDCap and informed electronic consent was obtained from all participants ^28^. Recruitment was done through social media, word of mouth, COVID-19 registries, presentations, as well as from Stanford HealthCare. We recruited participants with a confirmed or suspected COVID-19 infection, as well as those at high risk of exposure to COVID-19 (for e.g. via family members or patients), individuals with unknown respiratory illnesses, and individuals who did not report any illness. Participants were asked to wear their fitness tracker daily, as much as possible, and to download a study app called MyPHD (see Methods for Wearables Data Collection below) to share their wearables data.

### Metadata collection and surveys

Study metadata such as demographic information, reports of past illnesses, daily symptom tracking etc. were collected via REDCap. At enrollment, participants were asked to provide (i) demographic information such as age, sex, ethnicity, height, weight (ii) medical history including chronic illnesses, routinely taken medications etc., and (iii) COVID-19 illness status: whether they had a confirmed or suspected COVID-19 infection, if tested, then the test date, results, and symptom onset date.

In addition, all participants were asked to complete a daily symptom tracking survey, which tracked symptoms experienced and their severity on a scale of 1 - 5 (mild, mild-to-moderate, moderate, severe, worst-possible), body temperature (if recorded), any new tests or diagnoses for COVID-19 or other respiratory illnesses, test results, recovery dates, etc. Finally, participants were also asked to fill out a one-time past illness survey, where they could report past sick periods (up to 5 illnesses total) since November 1st, 2019. The past illnesses survey recorded the length of the sick period, and other elements similar to the daily survey: diagnoses (if any) of COVID-19 or other respiratory illnesses, any symptoms they reported experiencing during this period, as well as body temperature, and symptom severity on a scale of 1 - 5.

For this study, we restricted our analyses to a dataset of 31 individuals for who reported a positive COVID-19 diagnosis, a diagnosis date and/or symptom onset date (usually both n = 26), and wearables data appropriate for the analyses. Five of these individuals also reported other non-COVID-19 respiratory infections, including four individuals who reported two other illnesses since October 2019. Information was verified directly with the participants when necessary.

### MyPHD App for Wearables Data Collection

After participants enrolled on REDCap, we provided them with MyPHD, a smartphone app developed by our study team to collect their wearables data in a de-identified and encrypted manner. The MyPHD app was made available to study participants for both Android and iOS platforms. For Fitbit watches, the data were accessed through the Fitbit APIs and for wearable devices with Apple Healthkit integration, we obtained the data via Healthkit. Data transfer was done from source to a HIPAA compliant Google Cloud Platform project in an encrypted form, then the data was decrypted for pre processing and analysis in a controlled access, secure environment.

### Wearables Devices and Data Types Collected

Participants wore Fitbit smartwatches, including different versions such as Fitbit Ionic, Charge 4, Charge 3 etc. Data types collected included heart rate (HR), steps, and sleep. Raw heart rate, steps and sleep data were collected in JSON format. HR data was retrieved at 15-second resolution, steps values at a resolution of 1-minute, and sleep data as sleep stages intervals (wake, light, deep, and REM).

### Wearables Data Pre-processing

The retrieved raw HR/sleep/steps data from Fitbit were processed and integrated using a systematic workflow to produce a uniform format among different retrieval protocols. First, HR outliers (HR>200 and HR<30) were removed, as were all duplicates in the HR/steps/sleep data. Timestamps were unified to a standard timezone to be able to match different types of wearables data with metadata. HR features were extracted, such as median HR per minute, average HR per minute, change of variation of HR per minute, night-time resting heart rate, etc. Additionally, daily steps were calculated. For sleep features, total sleep duration per night and wake/light/deep/REM stages duration and their corresponding percentage for each night were calculated.

### Symptoms/Other Metadata Processing

Participant metadata and symptom surveys were downloaded and processed using a custom R and Python script. A total of 136 participants reported a positive COVID-19 diagnosis, but many were lacking a clear diagnosis or symptom date, or appropriate wearable data for the analyses. Height was converted to centimeters (Table 1), weight was converted to kilograms (Table 1), and reported temperature was converted to Fahrenheit (Fig. 4) for all participants.

### RHR-Diff offline detection

The resting heart rates (RHR) were obtained in the same approach as in Li et al. ^5^. For each person the RHRs were then standardized in one hour resolution based on the average of daily curves from a 28-day sliding window. The missing values in the RHRs were imputed as zeroes before the detection. We applied anomaly time interval detection based on rank scans from the work of Arias-Castro et al. ^14^ on the standardized residuals. Under significance level 0.05, the detected elevated time intervals were reported. To reduce possible false positives, short detected intervals less than 24 hour were removed.

### Anomaly Detection (offline)

HROS-AD (Heart Rate Over Steps Anomaly Detection) is an unsupervised anomaly detection model consisting of two major steps.

#### 1. Data pre-processing

We combine heart rate and step data from each user to compute a new feature known as HROS. HROS^(i)^ is the feature of the user *i*’s heart rate over steps (a value 1 is added to all steps to avoid the zero-division problem). Next, we used moving-averages (mean = 400 hours) and down-sampling (mean = 1 hour) to smoothen the time-series data and standardize further with Z-score transformation.

#### 2. Anomaly detection

When a HROS (Heat rate Over Steps) data point deviates markedly from others in a sample, it is called an anomaly or outlier. Any other expected observation is labeled as inlier.

We used *covariance.EllipticEvelope* class from *Scikit-learn* package ^15,16,29^ to fit a Gaussian distribution of the data, pointing out the anomalies that might be contaminating our dataset because they are extreme points in the general distribution of the dataset. For simplicity, we call this method as HROS-AD (Heart Rate Over Steps Anomaly Detector) if the input data is HROS. In HROS-AD, *EllipticEnvelope* is a function calculates the distance of each HROS observation with respect to the grand mean that takes into account all the observations in the data, and detect both univariate and multivariate outliers.

HROS-AD uses a key parameter called contamination, which can take a value up to 0.5 (Supplementary Table 22), which provides information about the proportion of the HROS outliers present in each dataset. We start with a value of 0.01 since it is the percentage of observations that should fall over the absolute value 3 in Z score distance from the mean in a standardized Gaussian distribution. If we don’t detect any anomalies, we gradually increase the contamination value from 0.01 until we find one. If we find too many anomalies with a 0.01 contamination score, we gradually decrease the contamination value. The predictions contain a vector of values 1 and −1 (1 being normal and −1 being anomalous).

We deleted the predictions if they were overlapping daytime (6AM to 12 AM) missing steps in the alert window of 21 days prior to the symptom and 7 days post symptom (21-7). There were 5 participants who had missing step data in at least one day in the 21-7 window and 3 of the participants had at least one prediction overlapping day time missing steps in the 21-7 window.

We also used the resting heart rate instead of HROS to check the model performance. We call this RHR-AD (Resting Heart Rate Anomaly Detector). It uses the same pipeline as HROS-AD except the input is resting heart rate. Resting heart rate was calculated as heart rate where steps are 0 for 10 mins ahead. We discarded 4 datasets that did not have enough data to train the model. Also, 2 of the participants had at least one prediction overlapping day time missing steps in the 21-7 window. Overall, the results between HROS-AD and RHR-AD were very similar.

### Activity and Sleep analysis

In our analysis, we only considered subjects that our RHR-diff algorithm detected change in their RHR any day between −14 pre symptoms date and 2 days after. We also removed subjects with more than 50% missing steps or sleep (each individually) in a window of −21 pre symptoms date and 7 days after. Following this filtration criteria, missing values were imputed using the last observation carried forward (LOCF) method. Afterward, daily steps and total sleep duration were Z-normalized for each person independently.

Since wearables sometimes have missing values (especially sleep, since some participants did not wear the watch every night), we evaluated the change in daily steps and total sleep duration without imputing the missing values. In a separate analysis (Fig. S4), we compared daily steps and total sleep pre/post-detection without the imputation process. We only considered 7 days pre-detection and 7 days post-detection.

Linear mixed models were conducted for daily steps and total sleep duration using the “nlme” package (version 3.1-142) in R. In our model, we included day annotation as a fixed effect and subject ID as a random effect. An anova test is applied on the fitted model to retrieve a p-value for the tested hypothesis.

### CuSum online detection

Based on the standardized residuals by comparing to the sliding window, CuSum statistics were calculated based on the work of ^20^. The values of CuSum statistics in the previous baseline days construct the null distribution. For the short-term data, 28 days were set as the baseline window and for the long-term data, 56 days were set. The missing values were removed. The threshold parameter in the CuSum statistic was set as the half of the 90%-quantile of the baseline residuals. Under the significance level 0.01, an alarm candidate is recorded when the first time that the CuSum statistic is significantly higher than the values from the null. We tracked the records of CuSum statistics for 48 hours. To reduce possible false positives, we started monitoring the statistic when it is above the threshold in the second hour. If within 24 hours the CuSum statistics stop increasing or within 48 hours the statistics are back to zero, the initial alarm will be removed.

### Visualization methods

We used ggplot2, Matplotlib, and matlab for plotting the most of the figures ^30,31^

## Data Availability

De-identified raw heart rate, steps, and sleep data used in this study can be downloaded from: https://storage.googleapis.com/gbsc-gcp-project-ipop_public/COVID-19/COVID-19-Wearables.zip

Data used for analyses and participant metadata is provided in supplemental tables.

https://storage.googleapis.com/gbsc-gcp-project-ipop_public/COVID-19/COVID-19-Wearables.zip

## Data availability

De-identified raw heart rate, steps, and sleep data used in this study can be downloaded from https://storage.googleapis.com/gbsc-gcp-project-ipop_public/COVID-19/COVID-19-Wearables.zip

## Acknowledgements

This work was supported by NIH grants, and gifts from the Flu Lab, and departmental funding from the Stanford Genetics department. We thank Diana Berrent from Survivor Corps for assistance in recruitment as well as Amy McDonough and Taylor Helgren from Fitbit Inc. for help in accessing Fitbit data. We thank Fitbit for promoting this study and for the donation of devices. The Stanford Healthcare Innovation Lab gratefully acknowledges the support of Alexandra Duisberg. Google Cloud Platform (GCP) costs were covered by Google for Education Academic Research and COVID-19 grant awards. This work was also supported by the Start-up/Departmental Funding and the HPC award for COVID-19 Research, which were both received by XL from Case Western Reserve University.

## Author Contributions

Conceptualization and study design: MPS, XL, TM

Project Supervision: XL, MPS

Project Administration: TM

IRB Review, Participant Recruitment & Coordination: AC, EH, OD-R, TM, BF, SK, MPS

Survey Data Collection: TM, AWB, AC, EH, OD-R, BF, SK, MG, RK

Wearables Data Collection & Processing: AAM, AB, AA

Data Coordination & Submission: AB, AA, AAM, AWB, TM

Software Development (MyPHD App): AB, AA

Algorithm Development: GKB, MW, XL

Data Analysis: TM, MW, AAM, GKB, MW, AWB, XL

Manuscript Preparation: TM, MW, AAM, GKB, AWB, XL, MPS

Manuscript Review and Editing: All co-authors

Funding: BR, XL, MPS

## Supplemental Figure Legends

**Fig. S1: Device Types**. Distribution of devices used by participants in the cohort

**Fig. S2: RHR-Diff and CuSum**. Examples of offline and online prediction using RHR-Diff for all 31 participants. For each pair of plots, the X-axis is the day (each tick is a day), and the Y-axis is either standardized heart rate residual (top panel), or CuSum statistic (bottom panel) for each participant. Red and purple vertical dashed lines indicate day of symptom onset and diagnosis respectively, for both panels. The green dashed line is at zero. Yellow/gold vertical solid lines mark the infection detection window used to score detections as a hit or a miss. Also indicated are time intervals where the standardized heart rate residuals are significantly elevated by RHR-Diff (red arrows in top panel), and alarm times from CuSum (blue vertical dashed lines).

**Fig. S3: HROS-AD plots**. Examples of anomalies detected by HROS-AD during COVID-19 infection for all 31 participants. For each panel, the X-axis is the number of days (each tick is a day), and the Y-axis is the standardized HROS (solid dark blue line). Red and purple vertical dashed lines indicate day of symptom onset and diagnosis, respectively. Detected anomalies are indicated by red dots.

**Fig. S4: Steps and Sleep Boxplots**. Boxplot showing changes in daily steps and total sleep duration between 7 days before and after the RHR-diff detection. Data presented in this figure represent days with sleep and steps data without imputing any missing data.

**Fig. S5: CuSum results for participants with long-term data**. Examples of CuSum results for four participants for whom long-term wearable data was available. For each participant, the X-axis is the day (each tick is a day), and the Y-axis is the CuSum statistic. Red and purple vertical dashed lines indicate day of symptom onset and diagnosis respectively, for both panels. Solid gold vertical solid lines mark the infection detection window used to score detections as a hit or a miss. Alarm times from CuSum are also indicated (blue vertical dashed lines).

## Listing of Supplemental Tables

**S1 Demographics and Health Characteristics of the Analyzed Cohort of 31 Individuals With COVID-19 Infection**

**S2 COVID-19 diagnoses for 31 participants**

**S3 Self-reported past illnesses S4 Daily Symptom Reports**

**S5 RHR-Diff Offline All Detected Intervals S6 HROS-AD Offline All Detected Anomalies S7 RHR-AD Offline All Detected Anomalies**

**S8 RHR-Diff COVID-19 Detection Timing and Standardized RHR Residuals S9 HROS-AD COVID-19 Detection Timing and HROS**

**S10 RHR-AD COVID-19 Detection Timing**

**S11 RHR-Diff Detection Timing w**.**r**.**t. Symptom Onset and Diagnosis Date**

**S12 RHR-Diff Delta HR in hourly windows during significant intervals around COVID-19 infection**

**S13 Standardized daily step count relative to illness onset for each participant**

**S14 Daily steps around detection day, in the infection window −21 days to +7 days of infection onset**

**S15 Daily steps +/− 7 days around the infection detection window**

**S16 Standardized total sleep duration relative to illness onset for each participant S17 Total sleep duration around detection day, in the infection window −21 days to +7 days of infection onset**

**S18 Total sleep duration +/− 7 days around the infection detection window S19 Online Detection Results Short-Term Data**

**S20 Online Detection Results Long-Term Data**

**S21 Comparison of RHR-Diff (Offline)and Online Detection Results**

## References

1. Sethuraman, N., Jeremiah, S. S. & Ryo, A. Interpreting Diagnostic Tests for SARS-CoV-2. JAMA (2020) doi:10.1001/jama.2020.8259.

2. Marinsek, N. et al. Measuring COVID-19 and Influenza in the Real World via Person-Generated Health Data. doi:10.1101/2020.05.28.20115964.

3. Dunn, J., Runge, R. & Snyder, M. Wearables and the medical revolution. Per. Med. 15, 429–448 (2018).

4. Kellogg, R. A., Dunn, J. & Snyder, M. P. Personal Omics for Precision Health. Circ. Res. 122, 1169–1171 (2018).

5. Li, X. et al. Digital Health: Tracking Physiomes and Activity Using Wearable Biosensors Reveals Useful Health-Related Information. PLoS Biol. 15, e2001402 (2017).

6. Perez, M. V. et al. Large-Scale Assessment of a Smartwatch to Identify Atrial Fibrillation. N. Engl. J. Med. 381, 1909–1917 (2019).

7. Radin, J. M., Wineinger, N. E., Topol, E. J. & Steinhubl, S. R. Harnessing wearable device data to improve state-level real-time surveillance of influenza-like illness in the USA: a population-based study. The Lancet Digital Health 2, e85–e93 (2020).

8. Zhu, G. et al. Learning from Large-Scale Wearable Device Data for Predicting Epidemics Trend of COVID-19. Discrete Dyn. Nat. Soc. 2020, (2020).

9. WHO Coronavirus Disease (COVID-19) Dashboard. https://covid19.who.int/.

10. Seshadri, D. R. et al. Wearable Sensors for COVID-19: A Call to Action to Harness Our Digital Infrastructure for Remote Patient Monitoring and Virtual Assessments. Frontiers in Digital Health vol. 2 (2020).

11. Arons, M. M. et al. Presymptomatic SARS-CoV-2 Infections and Transmission in a Skilled Nursing Facility. N. Engl. J. Med. 382, 2081–2090 (2020).

12. He, X. et al. Temporal dynamics in viral shedding and transmissibility of COVID-19. Nat. Med. 26, 672–675 (2020).

13. Witt, D., Kellogg, R., Snyder, M. & Dunn, J. Windows Into Human Health Through Wearables Data Analytics. Curr Opin Biomed Eng 9, 28–46 (2019).

14. Arias-Castro, E., Castro, R. M., Tánczos, E. & Wang, M. Distribution-Free Detection of Structured Anomalies: Permutation and Rank-Based Scans. Journal of the American Statistical Association vol. 113 789–801 (2018).

15. Rousseeuw, P. J. & Van Driessen, K. A Fast Algorithm for the Minimum Covariance Determinant Estimator. Technometrics vol. 41 212–223 (1999).

16. Garreta, R. & Moncecchi, G. Learning scikit-learn: Machine Learning in Python. (Packt Publishing Ltd, 2013).

17. Bi, Q. et al. Epidemiology and transmission of COVID-19 in 391 cases and 1286 of their close contacts in Shenzhen, China: a retrospective cohort study. The Lancet Infectious Diseases (2020) doi:10.1016/s1473-3099(20)30287-5.

18. Arias-Castro, E. & Wang, M. Distribution-free tests for sparse heterogeneous mixtures. TEST vol. 26 71–94 (2017).

19. Page, E. S. Cumulative Sum Charts. Technometrics vol. 3 1–9 (1961).

20. Levin, B. & Kline, J. The cusum test of homogeneity with an application in spontaneous abortion epidemiology. Stat. Med. 4, 469–488 (1985).

21. Mathew, D. et al. Deep immune profiling of COVID-19 patients reveals patient heterogeneity and distinct immunotypes with implications for therapeutic interventions. bioRxiv (2020) doi:10.1101/2020.05.20.106401.

22. Siordia, J. A. Epidemiology and clinical features of COVID-19: A review of current literature. Journal of Clinical Virology vol. 127 104357 (2020).

23. Xie, J. et al. Association Between Hypoxemia and Mortality in Patients With COVID-19. Mayo Clin. Proc. 95, 1138–1147 (2020).

24. Jouffroy, R., Jost, D. & Prunet, B. Prehospital pulse oximetry: a red flag for early detection of silent hypoxemia in COVID-19 patients. Critical Care vol. 24 (2020).

25. Lorente-Ros, A. et al. Myocardial injury determination improves risk stratification and predicts mortality in COVID-19 patients. Cardiol. J. (2020) doi:10.5603/CJ.a2020.0089.

26. Chen, R. et al. Personal omics profiling reveals dynamic molecular and medical phenotypes. Cell 148, 1293–1307 (2012).

27. Miller, D. J. et al. Analyzing changes in respiratory rate to predict the risk of COVID-19 infection. doi:10.1101/2020.06.18.20131417.

28. Harris, P. A. et al. Research electronic data capture (REDCap)--a metadata-driven methodology and workflow process for providing translational research informatics support. J. Biomed. Inform. 42, 377–381 (2009).

29. Boschetti, A. & Massaron, L. Python Data Science Essentials. (Packt Publishing Ltd, 2015).

30. Wickham, H. ggplot2: Elegant Graphics for Data Analysis. (Springer, 2016).

31. Hunter, J. D. Matplotlib: A 2D Graphics Environment. Comput. Sci. Eng. 9, 90–95 (2007).

